# Thalamocortical structural connectivity in children with focal epilepsy: a diffusion MRI, case-control study

**DOI:** 10.1101/2025.09.27.25336801

**Authors:** Rory J. Piper, Xiyu Feng, Aswin Chari, Maria H. Eriksson, Konrad Wagstyl, Kiran Seunarine, Gerald Hall, Peter N. Taylor, Yujiang Wang, Jonathan D. Clayden, Chris A. Clark, M. Zubair Tahir, David W. Carmichael, Torsten Baldeweg, Martin M. Tisdall

**Affiliations:** Developmental Neurosciences, UCL Great Ormond Street Institute of Child Health, University College London, London, UK; Department of Neurosurgery, Great Ormond Street Hospital, London, UK; School of Biomedical Engineering & Imaging Sciences, King’s College London, London, United Kingdom; Developmental Imaging and Biophysics Unit, Great Ormond Street Institute of Child Health, University College London, London WC1N 1EH, UK; School of Computing, Newcastle University, Newcastle upon Tyne, UK

**Author notes:** **Correspondence to:** Dr. Rory J. Piper. Developmental Neurosciences, UCL Great Ormond Street Institute of Child Health, University College London, 30 Guilford Street, London, UK, WC1N 1EH.

**Keywords:** thalamus, diffusion MRI, tractography, connectivity, epilepsy, children

## Abstract

**Objectives:** Determining patient-specific thalamic connectivity alterations may be an important step towards personalized surgical and neuromodulation strategies, but no data are available to support this concept in pediatric cohorts. This study investigated thalamocortical structural connectivity profiles in children with focal-onset epilepsy of different seizure onset zones.

**Methods:** This neuroimaging, case-control study compared structural connectivity of four thalamic nuclei (anteroventral (AV), centromedian (CM), mediodorsal (MDPf) and pulvinar (PUL)) between 81 children who underwent surgery for focal-onset epilepsy (median age=12.2 years) and 63 controls (median age=12.8 years). Using preoperative 3-tesla diffusion MRI, brain (Lausanne) and thalamic (THOMAS) parcellations combined with tractography generated structural connectomes based on streamline counts. Connectivity strength of each thalamic nucleus was calculated by summing the weights of each connecting brain region.

**Results:** Patients had higher structural connectivity strengths of the thalamic nuclei than controls (effect size (η²LJ)=0.146; p<0.001), differentially involving nucleus regions, but there was no overall difference in nucleus volumes (η²LJ<0.000; p=0.968). When comparing patient groups defined by seizure onset zones, it emerged that reduced AV connectivity strength was specific to the hippocampal sclerosis group, whereas CM, MDPf and PUL connectivity was similarly high in all the patient groups, including those with frontal or temporal lobe epilepsy. Patients who were seizure free after surgery had a lower ipsilateral and a higher contralateral connectivity strength (η²LJ=0.109; p=0.006) and volumes (η²LJ=0.073; p=0.025) of thalamic nuclei compared to those who were not.

**Significance:** This study provides unique data suggesting that different pediatric focal epilepsies have distinct structural thalamocortical connectivity and volumetric profiles. The structural connectivity and volumetric asymmetries of the thalami have an association with postoperative seizure freedom. More studies are required to further understand the thalamic connectivity signatures that may have implications for precision surgical planning and neuromodulation targeting for focal-onset epilepsy.

**Key points:**

- Children with focal epilepsy show overall higher thalamic structural connectivity than controls.
- Reduced structural connectivity of the AV is specific to children with hippocampal sclerosis.
- Patients who were seizure free post-surgery had lower ipsilateral but higher contralateral thalamic structural connectivity and volumes.
- Distinct thalamic connectivity patterns may guide personalized surgery and neuromodulation strategies in pediatric epilepsy.

## Introduction

Epilepsy is a neurological disorder that predisposes affected individuals to recurrent epileptic seizures^1^ and around one third of patients have *‘drug-resistant epilepsy’*^2,3^. Selected patients with focal-onset seizures may benefit from surgical resection and a subset becomes seizure free postoperatively^4^. However, despite decades of research and technological advancement within epilepsy surgery, the postoperative rate of seizure freedom remains at approximately 60%^5^ and further research is required to provide both data-driven methods to guide candidacy, predict postoperative outcomes^6^, and to develop alternative therapeutic approaches.

Epilepsy is now considered a brain network disorder^7^, and multiple studies have shown that patients with focal-onset epilepsy have abnormal brain networks that extend further than the putative seizure-onset zone^8,9^. The thalamus is increasingly being implicated as a key node of seizure propagation in the abnormal brain networks of patients with focal-onset epilepsies^10^. Diffusion MRI (dMRI) provides a method of investigating structural thalamocortical connectivity alterations and prior studies have shown reduced thalamocortical connections of the anterior (ANT) ^11^ and medial pulvinar (PUL)^12^ nuclei of the thalamus in adults with mesial temporal lobe epilepsy (TLE). Several studies have shown that thalamocortical network alterations on preoperative studies are associated with the seizure freedom outcomes following resective epilepsy surgery^13–15^. A dMRI study in a pediatric cohort of focal epilepsy of mixed etiology found group level bilateral network abnormalities in central, lateral and PUL nuclei^16^, highlighting a need to understand nucleus-specific alterations in different types of focal-onset epilepsy. Furthermore, data from volumetric MRI studies demonstrate that atrophy is evident in the thalamus in adult patients with TLE, particularly in the anterior, dorsomedial and pulvinar thalamic regions^11,17–19^, and higher in those with persistent post-operative seizures^19^.

Neuromodulation of the thalamus using deep brain stimulation (DBS) is gaining momentum as a therapeutic option in focal-onset epilepsy ^20,21^. The ANT, specifically the anteroventral (AV) nucleus of the ANT, is now an approved DBS target for treating focal-onset seizures in the USA and several European countries and is approved under certain conditions in the UK^22^. Other thalamic nuclei are being investigated as potential neuromodulation targets for epilepsy including the centromedian (CM), mediodorsal-parafasicular (MDPf) and PUL nuclei of the thalamus ^10^ ^23^. Selection of the optimal thalamic nucleus target for DBS in focal epilepsy may be individualized, depending on the seizure-onset zone and the specific pathological thalamocortical network^10^. It may be that invasive (stereo-EEG) and non-invasive (for example, dMRI) studies are able to refine thalamic targeting in DBS but a better understanding of thalamocortical networks in health and in different types of focal epilepsy is required.

Although there have been previous studies of thalamic structural connectivity, predominantly in the context of adult surgical TLE cohorts^11,18^, further studies are required in other focal epilepsies and in childhood cohorts. This study therefore investigates thalamocortical structural connectivity in children with focal epilepsy undergoing resective epilepsy surgery. This study compares connectivity with a cohort of healthy controls and explores differences in connectivity in children with different seizure-onset zones. The primary objectives were to investigate the structural connectivity profiles of four thalamic nuclei shown to have abnormal thalamocortical structural connectivity in prior adult focal epilepsy studies and those of interest as thalamic neuromodulation targets in epilepsy^23–25^. Secondary outcomes were 1) to investigate the association of these connectivity profiles with post-surgical seizure freedom, 2) to investigate these same questions using the volumes of the nuclei and 3) to investigate the relationship between the nuclei connectivity strength and volumes.

## Materials and methods

### Study design

This was a retrospective, neuroimaging, case-control study. Ethical approval for accessing the data was obtained locally by the UCL Great Ormond Street Institute for Child Health Research & Development Department (23NP01). This study has been performed in accordance with the standards of the *Helsinki Declaration*. Data from healthy participants were used from prior studies^26,27^. The reporting of this study adhered to the STROBE checklist.

### Cohorts

Imaging data were included for a pediatric cohort (aged 7-18 years) that underwent focal resective epilepsy surgery at Great Ormond Street Hospital between 2015 (implementation of a standardized diffusion MRI acquisition protocol) and 2023. Children younger than seven years of age were excluded to ensure that included patients were adequately age-matched to the control cohort (age range 7-18). All included children (patients and controls) had their imaging (preoperative for patients) acquired on the same MRI scanner with an identical protocol, as described below. Children were immediately excluded if they had undergone prior resections or had a confirmed diagnosis of Tuberous Sclerosis Complex.

Children with epilepsy were categorized into four seizure onset zone (SOZ) groups: (1) TLE with hippocampal sclerosis (HS) (TLE-HS); (2) TLE without HS (TLE-other); (3) frontal; and (4) other (including insular, parietal and occipital lobes). The TLE-HS was included to represent the ‘limbic’ epilepsy group and was confirmed using the postoperative histopathological report.

Neuroimaging data for healthy participants were available from prior studies performed at the UCL Great Ormond Street Institute of Child Health. To be included, healthy participants had to have had an identical neuroimaging protocol.

### Preoperative image acquisition

MRI data was acquired on a Siemens Magnetom Prisma 3T MRI at Great Ormond Street Hospital using a 20-channel head coil. MPRAGE images were acquired with a 1mm isotropic spatial resolution. Diffusion MRI data was acquired using a spin-echo single-shot 2D EPI acquisition and a multi-shell (b = 1000 & 2200s/mm^2^) and multiband (factor 2) sequence with 60 non-collinear diffusion directions, with 13 interleaved b = 0 images. The dMRI spatial resolution was 2mm in-plane with a 0.2mm gap across 66 slices. TRLJ=LJ3050LJms, TELJ=LJ60LJms, field of viewLJ=LJ220LJmmLJ×LJ220LJmm, matrix sizeLJ=LJ110LJ×LJ110, in-plane voxel resolutionLJ=LJ2.0LJmmLJ×LJ2.0LJmm, GRAPPA factor 2, phase-encoding (PE) partial FourierLJ=LJ6/8. An additional bLJ=LJ0 scan was acquired, with an identical readout to the diffusion-weighted scan, but with the phase encode direction flipped by 180° (in the anterior-posterior direction), for correction of susceptibility-related artefacts.

### Structural MRI processing

The MRI post-processing steps performed in this study used pre-existing and openly available research tools and is summarized in **Figure 1**. *Freesurfer (“recon-all”*; Version 7.2.3) was used to parcellate the brain in native space and manual control points were used to correct errors in intensity normalization^28^. *Freesurfer* parcellations were then converted to the *Lausanne parcellation* (*aparc125*) - chosen since it offers parcellation that was (a) higher-resolution and in the native space; and (b) anatomically accurate, and therefore surgically relevant.

**Figure 1.**
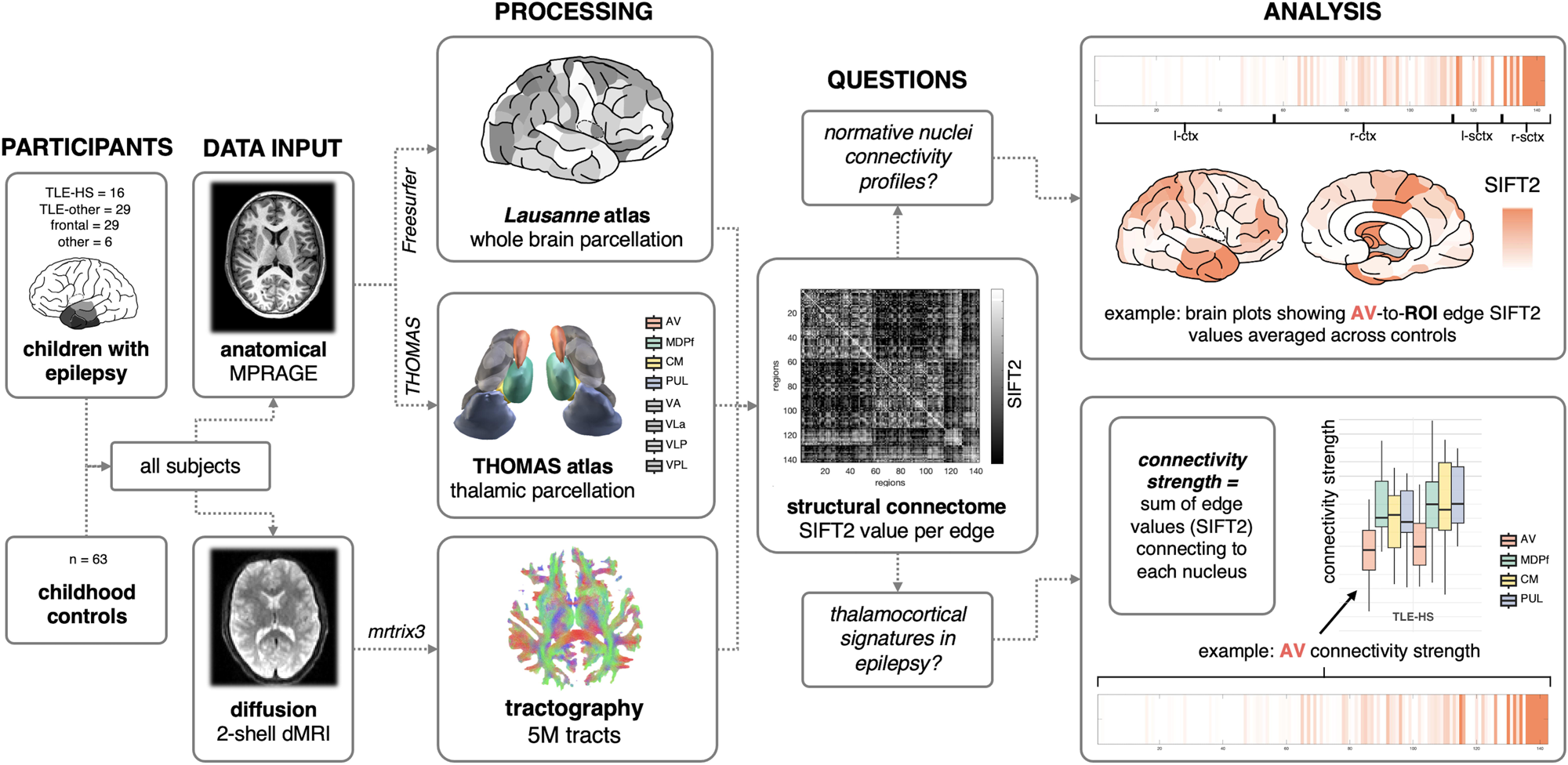
Image processing pipeline for the study. Image analysis pipeline from input data, connectome construction and analysis concepts. ctx = cortical regions; sctx = subcortical regions; TLE = temporal lobe epilepsy; TLE-HS = temporal lobe epilepsy with hippocampal sclerosis; TLE-other = temporal lobe epilepsy without hippocampal sclerosis; 5M = 5 million. AV = anteroventral nucleus of the thalamus; CM = centromedian nucleus of the thalamus; MDPf = mediodorsal-parafasicular nucleus of the thalamus; PUL = pulvinar nucleus of the thalamus.

The thalamic parcels of the Lausanne atlas were replaced with eight bilateral thalamic nuclei from the THOMAS atlas (AV = anteroventral, VA= ventral anterior; VLA = ventral lateral anterior; VLP = ventral lateral posterior; VPL = ventral posterolateral; PUL = pulvinar; CM = centromedian; MDPf = mediodorsal-parafasicular). The geniculate nuclei were excluded since not they were relevant and the habenular and mammillothalamic tracts were considered too small to utilize as seed regions for structural connectome analysis. The *T1w-THOMAS* software version was used, which segments the thalamic nuclei with an output in native space29.

Volumes of all parcels of the *THOMAS* atlas regions were extracted using MATLAB by summing the voxels in each parcel.

The estimated total intracranial volume was similarly determined using the derived value from *Freesurfer* (https://surfer.nmr.mgh.harvard.edu/fswiki/eTIV).

All imaging data were processed and analyzed in the native space to preserve accuracy and spatial resolution.

### Diffusion MRI processing & tractography

Diffusion MRI processing and tractography were performed using *MRtrix3*^30^. Data was denoized (*dwidenoise*^31,32^), corrected for inhomogeneity distortion (*dwifslpreproc*^33^), corrected for B1 field inhomogeneity (*FSL dwibiascorrect*^34^). Motion within the dMRI sequence was quantified for each subject by summing the displacement value measured between each direction (133), which is used later for regression. The T1w scan was rigidly registered to the diffusion scan using NiftyReg (*reg_aladin*^35^; http://cmictig.cs.ucl.ac.uk/wiki/index.php/NiftyReg) and the segmentations (*5ttgen fsl*^34^) resampled using the registration transforms (*reg_resample)*. Tractography was performed using *MRTRix3*scripts (*DWI2response dhollander*^36^, *DWI2fod* and five million streamlines were generated (Tckgen) and SIFT2^37^ assigned streamline weights to match estimated fibre densities in the underlying white matter (*Tcksift2*).

### Structural connectivity & statistical analyses

Whole brain structural connectomes were calculated per subject from tractography data using *tck2connectome* in *MRTRix3*. Each edge of the graph was the number of streamlines (using the *SIFT2* algorithm) between regions. Self-connections were omitted from the adjacency matrices*. Brain Connectivity Toolbox*^38^ (https://sites.google.com/site/bctnet) was used within *MATLAB* to calculate the connectivity *strength* of each thalamic nuclei – the sum of the edge *SIFT2* values / number of streamlines.

This study analyzed the connectivity of four bilateral thalamic nuclei from the *THOMAS atlas*, according to previous studies implicating their involvement in abnormal thalamocortical networks and currently explored thalamic neuromodulation targets in epilepsy: AV, PUL; CM and MDPF^23,25^.

Normative maps of the whole brain connectivity of the thalamic nuclei were generated by averaging the edge weights (*SIFT2* values) of the healthy control participants. The ipsilateral edges of the right and left nuclei were averaged (mean value) and represented on maps of the right hemisphere for visualization. The beta values from a general linear model were used to adjust the edge weights accounting for age, sex and motion: Normative maps are demonstrated using the *Simple Brain Plot MATLAB* function (https://github.com/dutchconnectomelab/Simple-Brain-Plot) ^39^. Since the map scale (**Figure 2**) is skewed by some very strong connections, the scale is limited to the 10^th^ and 90^th^ connectivity strengths.

**Figure 2.**
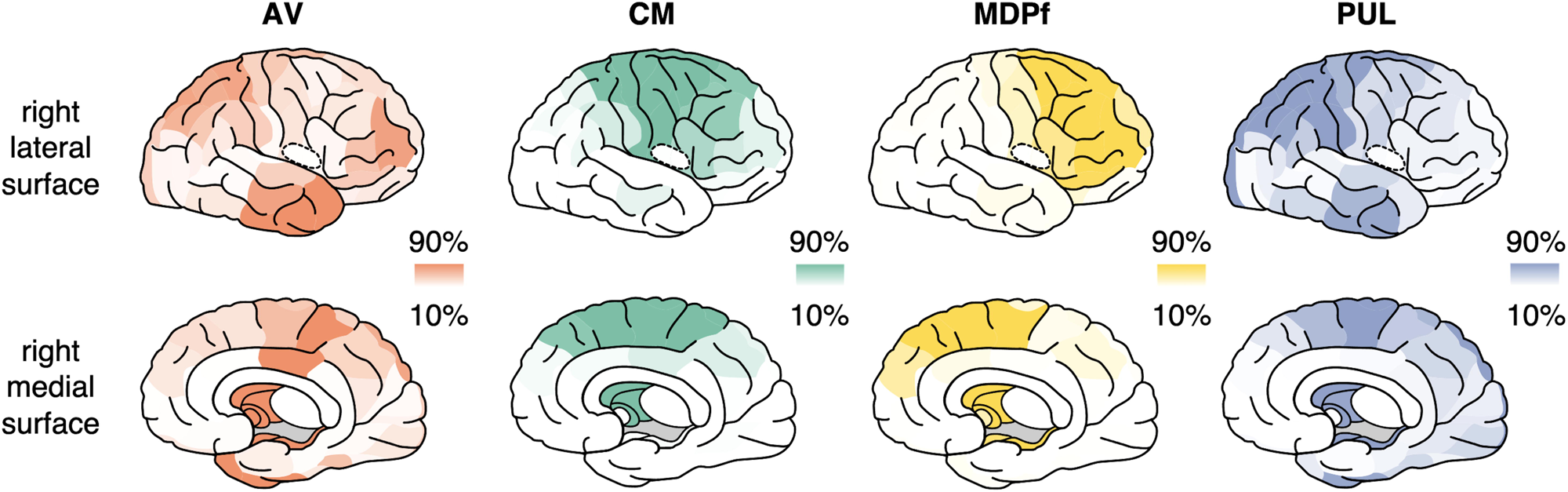
Structural connections of thalamic nuclei in children without epilepsy. Brain heat plots showing edge-wise structural connectivity values (SIFT2) between the AV (red), CM (centromedian), mediodorsal-parafasicular (MDPf) and pulvinar (PUL) nuclei with all other brain regions in the Lausanne Atlas (scale 2). The right and left values for each connection are averaged and demonstrated on the right hemisphere (lateral surface and medial surface). The scale bar for each nucleus includes the 10^th^ to 90^th^ percentile of the connection strengths.

Before analyzing thalamocortical strengths, the strength of each nucleus in the patient and control groups were z-scored against the distribution of the controls after using a general linear model (GLM) built using control data that accounted for age, sex, average ROI strength (mean of the connectivity strength of all the ROI across the whole brain parcellation, and total motion in dMRI sequence (aforementioned). To account for whole brain deviances in diffusion metrics, the mean ROI strength was entered into the GLM for strength z-scoring. Ipsilateral nuclei on the right side were z-scored to the corresponding right-sided nuclei in the control group, and vice versa. A similar GLM method was built for thalamic volumes, accounting for age, sex, and intracranial volume.

General linear models were used to model the structural connectivity scores and volumes of each of the thalamic nuclei, using the within-subjects factors *‘nucleus’* (AV, CM, MDPf, PUL), and *‘side’* (right or left), and between-subjects factors ‘group’ (controls and patients). Pillai’s trace was used to report the multivariate tests. Partial eta-squared effect sizes are provided, interpreted as small (η²LJ>0.01), moderate (η²LJ>0.06) or large (η²LJ>0.14) effects.

Another general linear model was used to model the structural connectivity strengths and volumes of these same thalamic nuclei between the patients with different seizure-onset zones: TLE-HS, TLE-other and frontal. Patients with other epilepsy localizations were not included in this subgroup analysis due to the small sample size and heterogeneity within this group. In addition to ‘nucleus’ the other within-subject factor included in the model was *‘laterality’* (ipsilateral, contralateral). *‘Postoperative seizure freedom’* (seizure free, not seizure free) was added as a second between-subjects factor.

Regional thalamocortical connectivity alterations were calculated by subtracting the control thalamocortical edges from the patient thalamocortical edges. Before the subtraction, a general linear model was created that inputted age, sex and motion to correct for these factors using the beta score. Right and left edges in patients were compared to the corresponding right and left edges in controls before the ipsilateral edges in patients were selected and projected on the right hemispheric visualizations, again using *Simple Brain Plot*.

Subject data were excluded from the analysis based on quality control checks of the scan motion artefact, automated parcellation, and/or tractography.

Overall summary data are presented as median values and IQR. **Figures 3** and **4** show the estimated means and one standard error of the mean. Statistical significance was set at p<0.05. Statistical testing was performed using R (Version 4.1.0) and SPSS (version 29).

**Figure 3.**
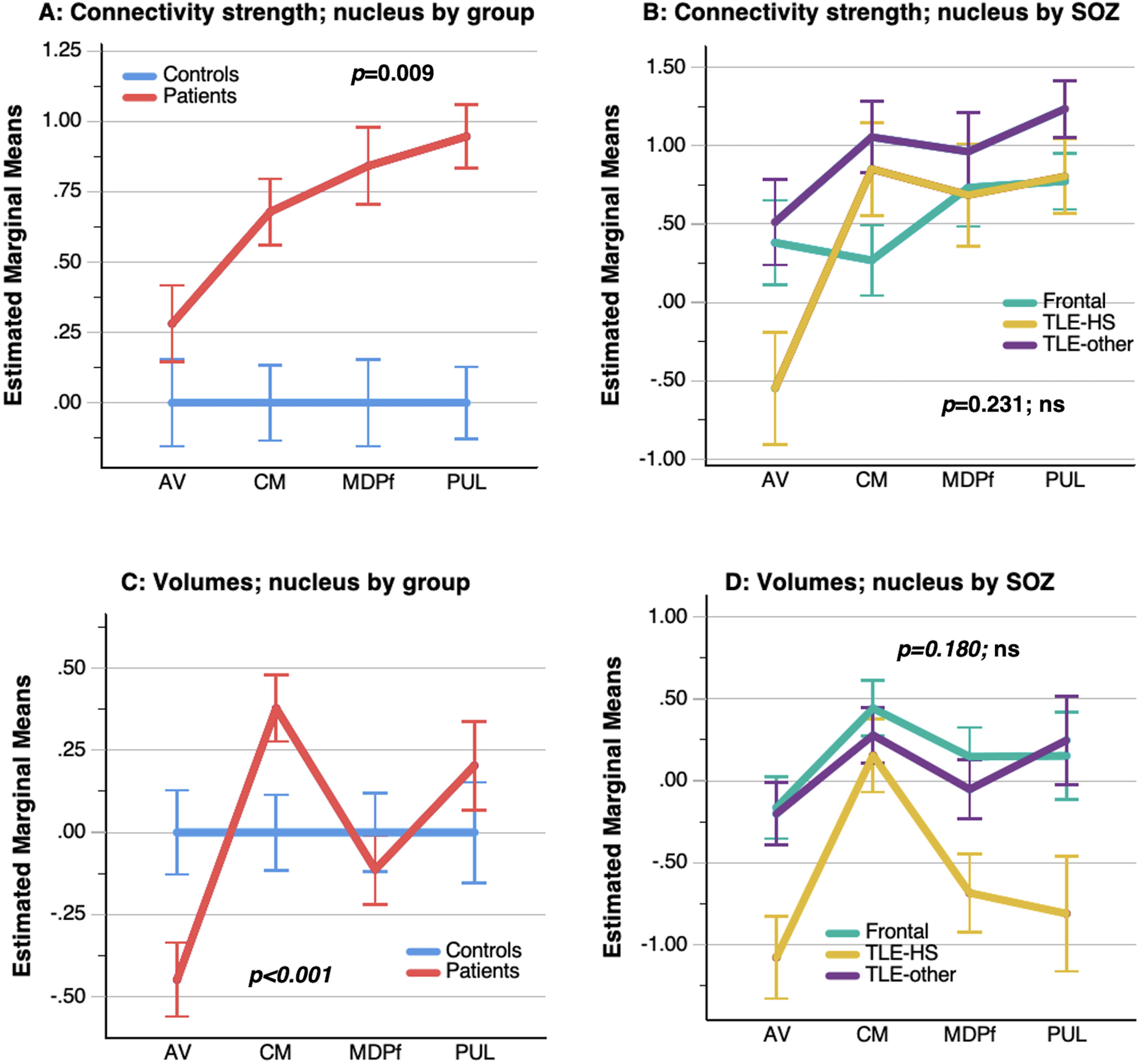
Estimated marginal means of the nucleus connectivity strengths and volumes (A & C, respectively), comparing the controls and patients and comparing the different focal epilepsy groups (B & D, respectively). The mean value is presented with one standard error of the mean shown by the whiskers. TLE-HS = temporal lobe epilepsy with hippocampal sclerosis; TLE-other = temporal lobe epilepsy without hippocampal sclerosis; Ipsi. = ipsilateral; Contra. = contralateral; AV = anteroventral nucleus of the thalamus; CM = centromedian nucleus of the thalamus; MDPf = mediodorsal-parafasicular nucleus of the thalamus; PUL = pulvinar nucleus of the thalamus.

**Figure 4.**
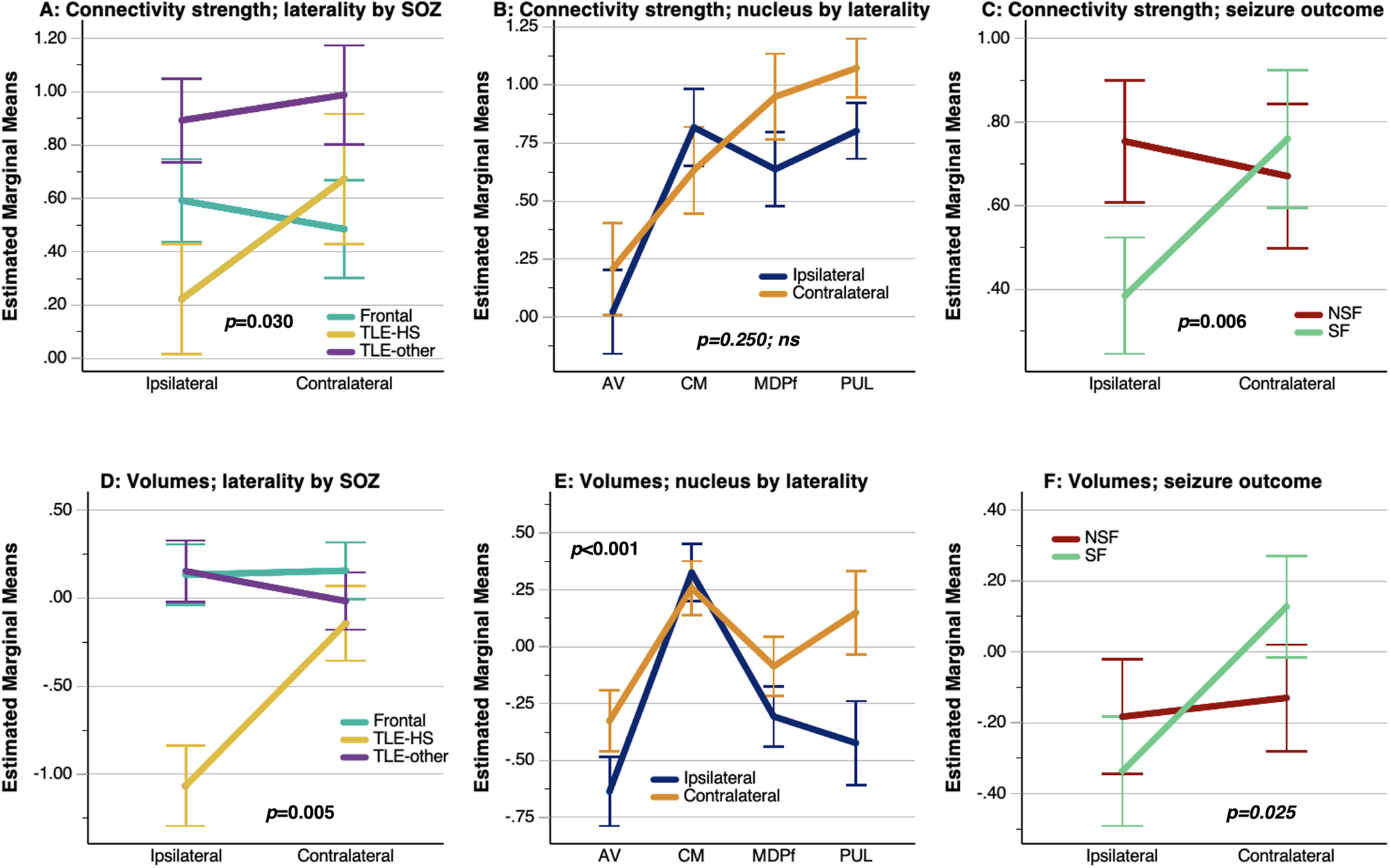
Estimated marginal means of the nucleus connectivity strengths (A-C) and volumes (D-F) in children with focal epilepsy. Comparison of ipsilateral and contralateral nucleus connectivity strength and volume in different focal epilepsy groups (**A & D**) and across different thalamic nuclei (**B & E**) and postoperative seizure outcomes (**C & F**), respectively. The mean value is presented with one standard error of the mean shown by the whiskers. TLE-HS = temporal lobe epilepsy with hippocampal sclerosis; TLE-other = temporal lobe epilepsy without hippocampal sclerosis; Ipsi. = ipsilateral; Contra. = contralateral; AV = anteroventral nucleus of the thalamus; CM = centromedian nucleus of the thalamus; MDPf = mediodorsal-parafasicular nucleus of the thalamus; PUL = pulvinar nucleus of the thalamus; NSF = not seizure free; SF = seizure free.

## Results

### Patient and control cohort characteristics

This study included 81 patients (median age 12.2 years; IQR 9.6–16.0 years; 40/81 female) who underwent surgical resection of a putative seizure-onset zone for drug-resistant focal-onset seizures. 45 patients had temporal resections (of whom 16/45 had HS), 29 had frontal resections, and 7 had other (insular, parietal, occipital or multi-lobar) resections. Tumours (24/81), focal cortical dysplasia (FCD) (23/81), and hippocampal sclerosis (HS) (16/81) were the most frequent pathologies. 47/81 (58%) were known to be seizure free at final clinical follow-up (median duration of follow up 1.7 (IQR 1.2-2.8) years). Two patients did not have postoperative seizure freedom data available. A comparison group of 63 healthy participants (median 12.8 years; IQR 9.6–14.5 years; 49/63 female) were included. Characteristics of the patient and control cohorts are provided in full in **Table 1**.

### Thalamocortical structural connectivity in healthy controls

Discounting the inter-thalamic nuclei connections, the strongest structural connections of the nuclei in healthy controls are projected in **Figure 2** and ranked in **Supplementary Table 1**. Raw (not adjusted for age, sex, or other variables) connectivity strength of the right (Pearson R=-0.37; p=0.01) and left (Pearson R=-0.35; p=0.01) MDPf and the right PUL (Pearson R=-0.31; p<0.05) decreased with age across the control cohort, but not in the patients. No significant trends were seen in the other nuclei in any of the groups **(Figure S1A)**.

### Thalamic nuclei connectivity signatures in focal epilepsy groups

Overall, thalamic nuclei connectivity strength was higher in patients than in controls (F=24.325; η²LJ=0.146; p<0.001), but this differed by nucleus region and was least pronounced for the AV (**Figure 3A**). Full output from the GLM model is provided in **Appendix 1**.

In another GLM investigating the nuclei connectivity strengths in the TLE-HS, TLE-other and frontal SOZ groups only, although there was no between-subjects effect of the SOZ group (F=2.327; η²LJ=0.065; p=0.105) or overall *group* by *nucleus* effect (F=1.370; η²LJ=0.059; p=0.231), only the AV showed reduced connectivity strength in TLE-HS group (**Figure 3B**). A *laterality* effect, with ipsilateral connectivity strength reduction, was observed in TLE-HS but not in the other groups (*group* by *laterality* effect (F=3.690; η²LJ=0.099; p=0.030) (**Figure 4A**). Finally, *postoperative seizure freedom* interacted with *laterality* (F=8.176; η²LJ=0.109; p=0.006) by showing an asymmetrical strength profile (ipsilateral reduction) in comparison with a more bilateral profile in the not seizure free group (**Figure 4C**). Full output from the GLM model is provided in **Appendix 3**.

An exploratory analysis of average regional thalamocortical (edge-wise) connectivity alterations in patients compared to controls is presented in **Figure 5**. Widespread reductions in connectivity of the AV were observed in the TLE-HS group, but not the other groups. A consistent feature of increased structural connectivity of the paracentral regions with the CM nucleus is seen across the epilepsy cohorts.

**Figure 5.**
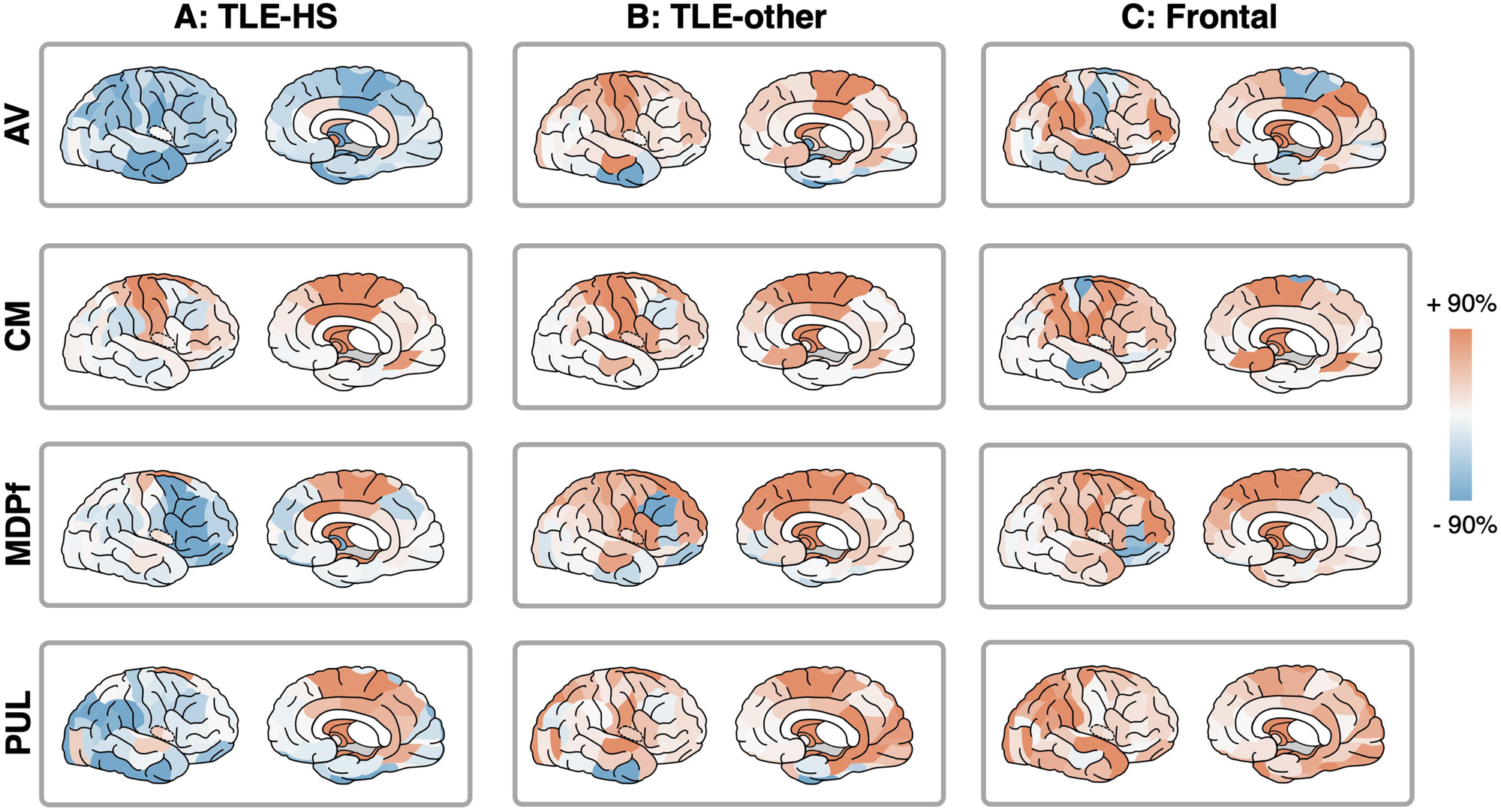
Difference in thalamocortical connectivity strengths between patient groups and healthy controls. The maps show ipsilateral nuclei-to-ROI connections visualized on the right hemisphere surfaces. Red color shows higher values in patients and blue colors show lower values in patients compared to controls. The scale bar for each nucleus includes the 10^th^ to 90^th^ percentile of the connection strengths.

### Volumes of thalamic nuclei in focal epilepsy sub-groups

For nuclei volume, there was no overall difference in volumes between patients and controls (F=0.002; η²LJ<0.000; *p*=0.968). A *nucleus* by *group* interaction was significant on multivariate testing (F=6.051; η²LJ=0.115; p<0.001), showing reduced volume of the AV and increased volume of the CM in patients compared to controls (**Figure 3C**). Full output from the GLM model is provided in **Appendix 2**.

In another GLM investigating the nuclei volumes in the TLE-HS, TLE-other and frontal groups only, the TLE-HS group showed a distinct reduction of AV, MDPf, PUL but not CM volumes compared to the other two groups (**Figure 3D**).

*Laterality* interacted with *SOZ* by showing ipsilateral reductions in TLE-HS but not the other groups (F=11.873; η²LJ=0.262; p<0.001) (**Figure 4D**). A further *laterality* with *nucleus* interaction (F=7.952; η²LJ=0.268; p<0.001) (**Figure 4E**) revealed that ipsilateral volume reductions were found in the AV, MDPf and PUL but not the CM. There was an interaction between *seizure freedom* and *laterality* (F=5.237; η²LJ=0.073; p=0.025), with the seizure free group showing an asymmetrical volume profile (ipsilateral reduction) in comparison with a more bilateral profile in the not seizure free group (**Figure 4F**). A marginal *nucleus* by *laterality* by *SOZ* interaction (F=2.245; η²LJ=0.093; p=0.043) was driven by reduced ipsilateral volumes in the TLE-HS group. Full output from the GLM model is provided in **Appendix 4**.

## Discussion

This was a retrospective, case-control, neuroimaging study analyzing the structural connectivity of four thalamic nuclei of the thalamus (AV, CM, MDPf and PUL), in a control sample of healthy developing children and in children with focal-onset seizures. Prior studies of thalamic connectivity have primarily focused on adult patients with TLE, and this study provides new insights into the thalamocortical structural connectivity in children in normal development and the alterations found in different focal epilepsies.

This study shows the novel findings that (a) overall thalamocortical structural connectivity is higher in children with focal epilepsy compared with controls; (b) TLE-HS has a distinct profile of thalamocortical connectivity, with a laterality effect (reduced ipsilateral connectivity), particularly of the AV nucleus, when compared to other focal epilepsy groups; and (c) there is a laterality effect associated with postoperative seizure freedom following resective surgery for children with focal epilepsy, with reduced connectivity strength of the ipsilateral compared to contralateral thalamic nuclei.

### *‘Normal’* thalamocortical connectivity in controls

This study demonstrates the anticipated structural connectivity profiles of the thalamic nuclei as previously described in the literature in a cohort of healthy controls (**Figure 2 & Table S1**). The AV nucleus has been reported to have strong connectivity with the limbic structures and is a critical node in the *Circuit of Papez*, with structural connectivity to the mammillary body (mammillothalamic tract), fornix, cingulate gyrus, retrosplenial cortex and mesial temporal lobe^40^. The CM nucleus is reported to have strong connectivity with the primary motor and sensory cortices and, although not studied in the brain parcellation used in this current study, the brainstem and cerebellum^41,42^. The mediodorsal nucleus (in this study combined with the parafasicular nucleus (MDPf)) has strong connectivity with the prefrontal cortices ^43–45^. A recent dMRI study has reported the PUL as having four distinct subregions and ‘fibre contingents’, including an anterior component with fibres extending to the anterior temporal lobe, a lateral component with fibres to the lateral temporal lobe, an ‘optic radiation-like’ component reaching the posterior basal temporal lobe, and an ‘arcuate fasciculus-like’ component extending to the temporal operculum^46^.

### Distinct thalamocortical profiles in focal epilepsy

In a general linear model, overall thalamic nuclei connectivity strength was found to be higher in patients with focal epilepsy when compared to controls. To our knowledge, this is not well documented in the prior literature, other than another study from our centre that showed higher weighted degree (effectively the same as the measure of ‘connectivity strength’ used in this study) of the pulvinar, central-lateral and lateral-posterior nuclei in children compared to controls^16^. Decreased connectivity of the AV, MDPf and PUL has been documented in adult TLE^11,18^, but there is no other data to conclude the structural connectivity profiles of thalamic nuclei in other forms of focal epilepsy. To ensure these high thalamic connectivity strength findings are not simply reflective of a whole brain connectivity strength effect, the mean node strength per patient is added to the general linear model before z-scoring thalamic values to controls. Furthermore, **Figure S3** shows a heterogeneous distribution of increased and decreased node strength across the brain when comparing the individual strength of each brain region between patient to controls.

This study shows that the TLE-HS group have a distinct profile of thalamocortical structural connectivity when compared to patients with TLE-other and frontal epilepsy (**Figure 3 & 4**). Although, overall, thalamic nuclei strength is high in patients compared to controls, this study also finds the TLE-HS group is predominantly characterized by lower AV connectivity strength values compared to controls and other focal epilepsy groups (**Figure 3B**). This finding matches the abnormalities in AV connectivity have been reported across several studies of adult patients. For example, Keller et al. found decreased streamline counts between the temporal lobe with the AV in patients with TLE-HS^18^. Yilirim et al. showed decreased structural connectivity of the AV with the hippocampus in adult patients with TLE-HS, but not in those with MRI-negative TLE^11^. Furthermore, an fMRI study by Vaughn et al. suggested increased clustering coefficient detected in the anterior thalamus in patients with TLE-HS, but not in patients with MRI negative TLE^47^.

Increased connectivity strength and volume of the CM nucleus was a consistent finding across all the epilepsy subgroups. On inspecting the whole brain connections (edges) of the nuclei (patients minus controls; **Figure 5**), a consistent feature is increased connectivity of the paracentral (motor and sensory) cortices. These features have not been reported in other studies of thalamocortical connectivity in focal epilepsy and needs corroboration. One speculation is the existence of an inhibitory network consistent across different epilepsy types^48^.

### Implications for epilepsy surgery

This study showed an asymmetry of the thalamocortical structural connectivity strengths in patients who were seizure free following resective epilepsy surgery (**Figure 4**). In patients who were seizure free after surgery, the ipsilateral nuclei strength and volume were lower than on the contralateral side, unlike those with recurring seizures who showed a more bilateral pattern. The reasons for this association are unclear, and no such finding is evident in the prior literature, however, a similar trend was observed here for thalamic volumes, akin to findings in adult TLE surgery^19^. Prior fMRI studies have shown the relevance of thalamic functional connectivity on post-surgical outcomes. For example, data from our own centre show that pediatric TLE patients who achieved seizure freedom after temporal lobe surgery exhibited stronger connectivity between the ipsilateral hippocampus and superior thalamus compared to those who were not seizure-free^49^. This may indicate a more localized ipsilateral propagation pathway, compared to a more distributed network in those not seizure free. In support, He et al found higher functional connectivity using resting-state fMRI (degree and eigenvector centrality) in the (entire) thalamus in adult patients with TLE who were not seizure free compared to those were seizure free following surgery, suggesting that ‘thalamic hubness’ could be a marker of a spatially wider or more complex network and a potential means of predicting risk of post-operative seizure recurrence^50^. From this study, we speculate that more localized or ipsilateral changes in connectivity could predict better surgical outcomes, but further data is required to investigate this.

### Implications for neuromodulation for epilepsy

Fisher et al.’s SANTE trial in 2010 treated patients with drug-resistant focal-onset seizures with bilateral ANT deep brain stimulation (DBS)^21^. The ANT was targeted regardless of the presumed seizure-onset zone and, although the study was not powered to assess this, the patients with temporal lobe onset seizures showed statistically significant benefits with ANT DBS whereas those with other epileptogenic foci did not. Given our updated knowledge since the SANTE trial, it seems increasingly unlikely that ANT stimulation is a one-size-fits-all approach to neurostimulation targeting for epilepsy DBS/RNS. Although the ANT is a favourable target to modulate the limbic network, it may not be as effective in treating non-mesial TLE epileptogenic networks. It may be that preoperative and non-invasive methods (such as diffusion MRI and tractography) can identify pathological networks specific to the individual or in patients with similar seizure-onset zones. Further studies are required to correlate thalamic connectivity alternations found on imaging with ground-truth data from thalamic stereo-EEG.

### Study strengths

A key strength of this study was its inclusion of a large sample of children with focal epilepsy, each with well-defined seizure onset zones and postoperative outcome data. This study overcame the limitations of prior studies that have predominantly focused on adult patients with only mesial TLE. The patients and controls had a high-quality, multi-shell diffusion MRI acquisition. The data demonstrates that motion during the dMRI sequence was not worse in patients, but in fact improved, largely due to many patients needing intubation for the scan (**Figure S2**). Importantly, this difference did not account for the group connectivity findings reported here.

Another strength of this study is the inclusion of the normal connections of the thalamic nucleus (**Figure 2 & Table 1**), which brings context to the differences found between patient and control data. Furthermore, although not the primary objective, the study identifies the normal and abnormal developmental trends in thalamic nucleus connectivity and volumetric data, in controls and patients, respectively (**Figure S1**). This study uses a GLM to account for these trends and to accordingly adjust the group-level results.

### Study limitations

This study had several limitations. Firstly, the measure of *connectivity strength* (the total of all nucleus-to-ROI weights) is an over-simplification of the thalamocortical network and does not account for the regional differences in connectivity strengths with the thalamic nuclei (demonstrated in **Figure 5**). Future work needs to better understand this variability and how this is clinically relevant. Furthermore, diffusion MRI and structural connectivity abnormalities may reflect the functional network abnormalities or epileptic network but are not the same. Diffusion MRI alterations may reflect the more chronic structural effects of an epileptogenic network and many only show interpretable changes in particular pathologies. For example, the TLE-HS thalamocortical abnormalities, particularly with reduced connectivity of the ANT, matches prior studies and is more easily explainable in terms of atrophy. In contrast, the other focal epilepsy groups did not show such obvious reductions in AV (or other nuclei) connectivity strength (**Figure 3C**).

### Future studies & next steps

Future works should aim to compare thalamocortical networks in both controls and patients with focal epilepsy across different imaging modalities. Comparison could be made between structure (volume and dMRI), and functional MRI, scalp EEG, and stereo-EEG that are more reflective of the real-time epileptic brain network. Furthermore, utilizing ultra-high-resolution (7-tesla) MRI may improve the accuracy of connectivity measures seeded from these small thalamic nuclei.

Further studies using thalamic stereo-EEG are, however, justified to provide patients with personalized neuromodulation strategies. Comparison of our findings to adult data, particularly in patient groups other than TLE-HS, would be interesting given that typically the duration of disease is longer. It may be that structural connectivity findings in those other focal epilepsy groups are more engrained and detectable in adult populations.

## Conclusions

This neuroimaging study provides unique insights to the thalamocortical structural connectivity and volumetric profiles of thalamic nuclei in children with and without focal epilepsy. Connectivity and volumetric thalamic profiles are distinct between children with different seizure-onset zones, which may have implications for the personalization of neuromodulation therapies such as DBS. The association of structural thalamic asymmetry with post-surgical seizure freedom may also suggest a potential role in improving selection of candidates for resective surgery.

## Acknowledgements

RJP was funded by the National Institute for Health Research (NHIR) & the Great Ormond Street Hospital Children’s Charity.

## Supporting material

Supporting documentation is available online.

## Author contributions

RJP conceptualized and designed the study, acquired the data, performed the analysis, wrote the first draft of the article and takes overall responsibility for the work. MHE acquired the data, interpreted the data and contributed to the writing of the article. XF, AC, MHE, KW, KS, GH, PNT, YW, JDC, CAC, MZT, DWC, TB, and MMT have interpreted the data and contributed to the writing of the article.

## Data availability

The shell scripts and MATLAB code to process the data are openly available: https://github.com/roryjpiper/thalamus_dMRI_epilepsy.

