## Supporting Information (Supplementary) for "Thalamocortical structural connectivity in children with focal epilepsy: a diffusion MRI, case-control study"

**Table S1.** **Strongest structural connections of each of the four studied thalamic nuclei (AV, CM, MDPf and PUL).** The strongest 10 connections are (determined by nucleus-to-ROI edge weight, SIFT2 value adjusted for age, sex, average strength in ROI nodes, total motion, and total intracranial volume) are ranked from strongest (rank 1) to weakest (rank 10). Inter-thalamic connections have been discounted. AV = anteroventral; CM = centromedian; MDPf = mediodorsal-parafasicular; ROI = region-of-interest; PUL = pulvinar.

| **Rank** |  | **AV-to-ROI strongest edges** | |  | **CM-to-ROI strongest edges** | |  | **MDPf-to-ROI strongest edges** | |  | **PUL-to-ROI strongest edges** | |
| --- | --- | --- | --- | --- | --- | --- | --- | --- | --- | --- | --- | --- |
|  |  | **ROI labels** | **Adjusted edge weights (SIFT2)** |  | **ROI labels** | **Adjusted edge weights (SIFT2)** |  | **ROI labels** | **Adjusted edge weights (SIFT2)** |  | **ROI labels** | **Adjusted edge weights (SIFT2)** |
| 1 |  | Caudate | 106254.56 |  | Caudate | 314006.12 |  | Caudate | 4754340.65 |  | Hippocampus | 15172300.64 |
| 2 |  | Hippocampus | 9434.96 |  | Superiorfrontal_4 | 40868.82 |  | Putamen | 304393.43 |  | Caudate | 4252873.55 |
| 3 |  | Putamen | 7171.74 |  | Precentral_1 | 16751.81 |  | Pallidum | 214128.73 |  | Putamen | 407385.64 |
| 4 |  | Pallidum | 6084.57 |  | Precentral_2 | 11348.05 |  | Superiorfrontal_4 | 65066.02 |  | Precentral_3 | 383096.67 |
| 5 |  | Accumbens-area | 5860.471482 |  | Postcentral_2 | 10856.94 |  | Superiorfrontal_3 | 53036.72 |  | Superiorparietal_3 | 333725.36 |
| 6 |  | Amygdala | 3087.70 |  | Superiorfrontal_3 | 10261.25 |  | Parstriangularis_1 | 44240.20 |  | Superiorparietal_1 | 260020.12 |
| 7 |  | Temporalpole_1 | 289.09 |  | Paracentral_1 | 9848.97 |  | Superiorfrontal_2 | 38187.90 |  | Superiorfrontal_4 | 258014.26 |
| 8 |  | Paracentral_1 | 272.27 |  | Pallidum | 9118.66 |  | Hippocampus | 32972.47 |  | Superiorparietal_2 | 233139.42 |
| 9 |  | Inferiortemporal_1 | 192.3295029 |  | Putamen | 8968.489122 |  | Rostralmiddlefrontal_1 | 31320.02603 |  | Inferiorparietal_2 | 218855.5076 |
| 10 |  | Posteriorcingulate_1 | 180.27 |  | Caudalmiddlefrontal_1 | 7416.38 |  | Rostralmiddlefrontal_2 | 30609.95 |  | Supramarginal_2 | 218130.62 |

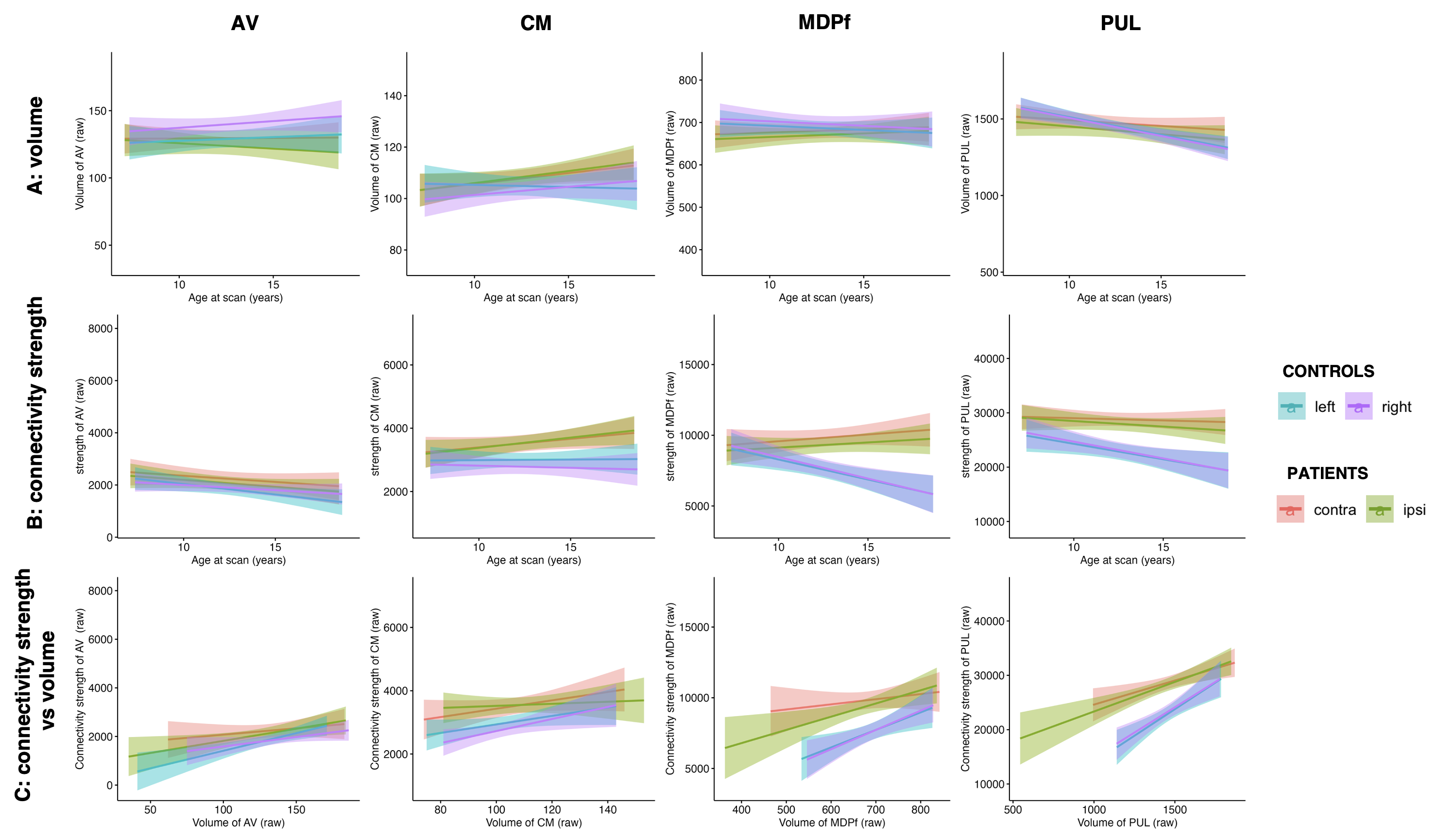

**Figure S1.** **Relationship between age, and nuclei connectivity strength and nuclei volume in each of the control (right and left) and patient (ipsilateral and contralateral) subjects.** (A) Pearson correlation between nuclei connectivity strength and age at scan. (B) Pearson orrelation between nuclei volume and age at scan. (C) Correlation between nuclei volume and nuclei connectivity strength. Raw (not adjusted for age, sex, or other variables) connectivity strength of the right (R=-0.37; p=0.01) and left (R=-0.35; p=0.01) MDPf and the right PUL (R=-0.31; p<0.05) decreased with age across the control cohort, but not in the patients. No significant trends were seen in the other nuclei in any of the groups Adjustment for multiple comparisons was performed using the Holm method.

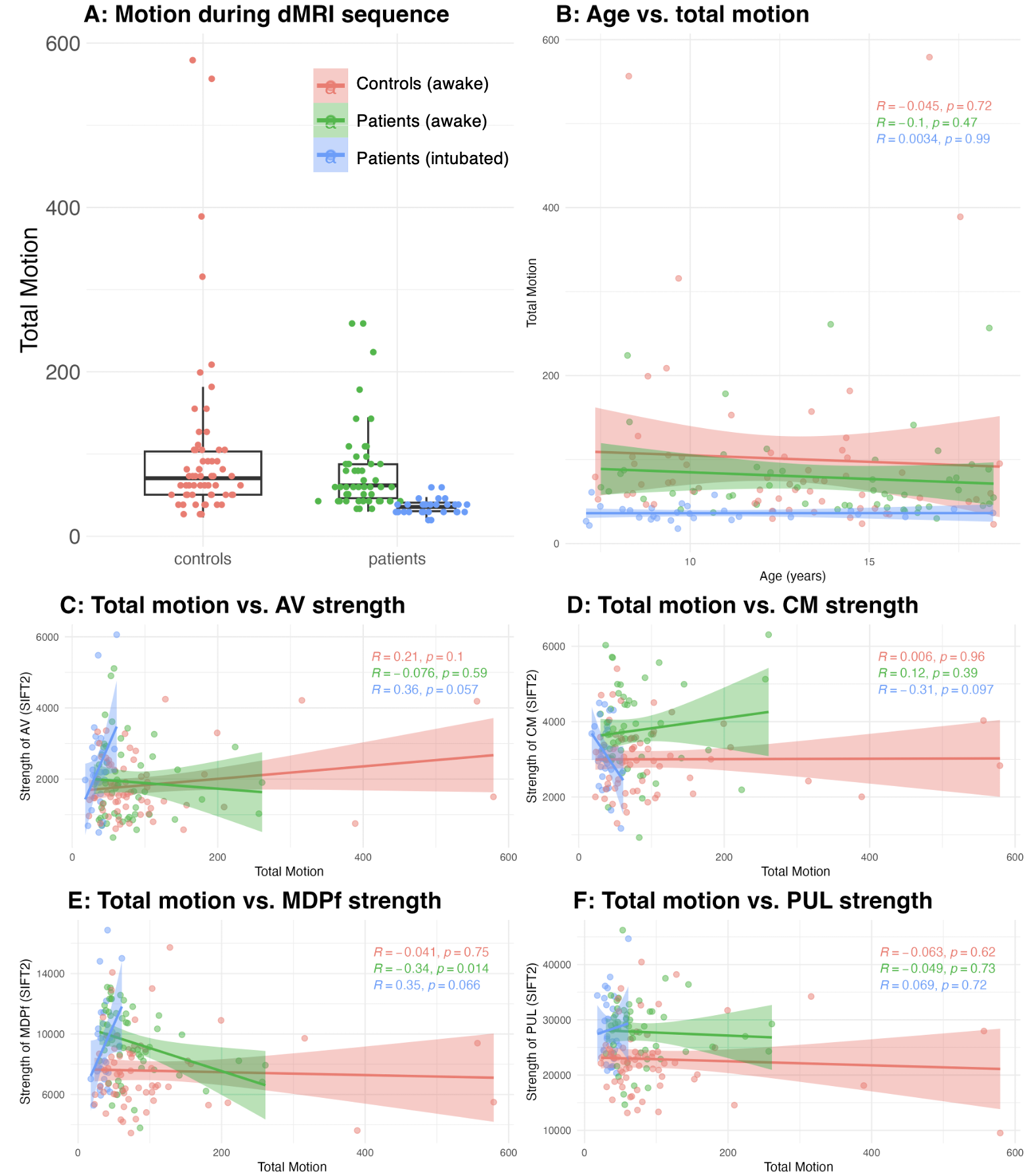

**Figure S2. Comparison of subject motion during the dMRI sequence between controls, patients and intubation status.** An estimation of motion is derived from the output from eddy in *MRtrix3* and summing the total deviation of the direction acquisitions compared to the first direction acquisition. An ANOVA was performed (motion ~ group * intubation) that found differences between the groups (F=8.405; p<0.01) and intubation status (F=5.728; p<0.05). **C-F** show the Pearson correlation between the total motion and the nuclei strength in each per group: controls awake (red), patients awake (green), patients intubated (blue).

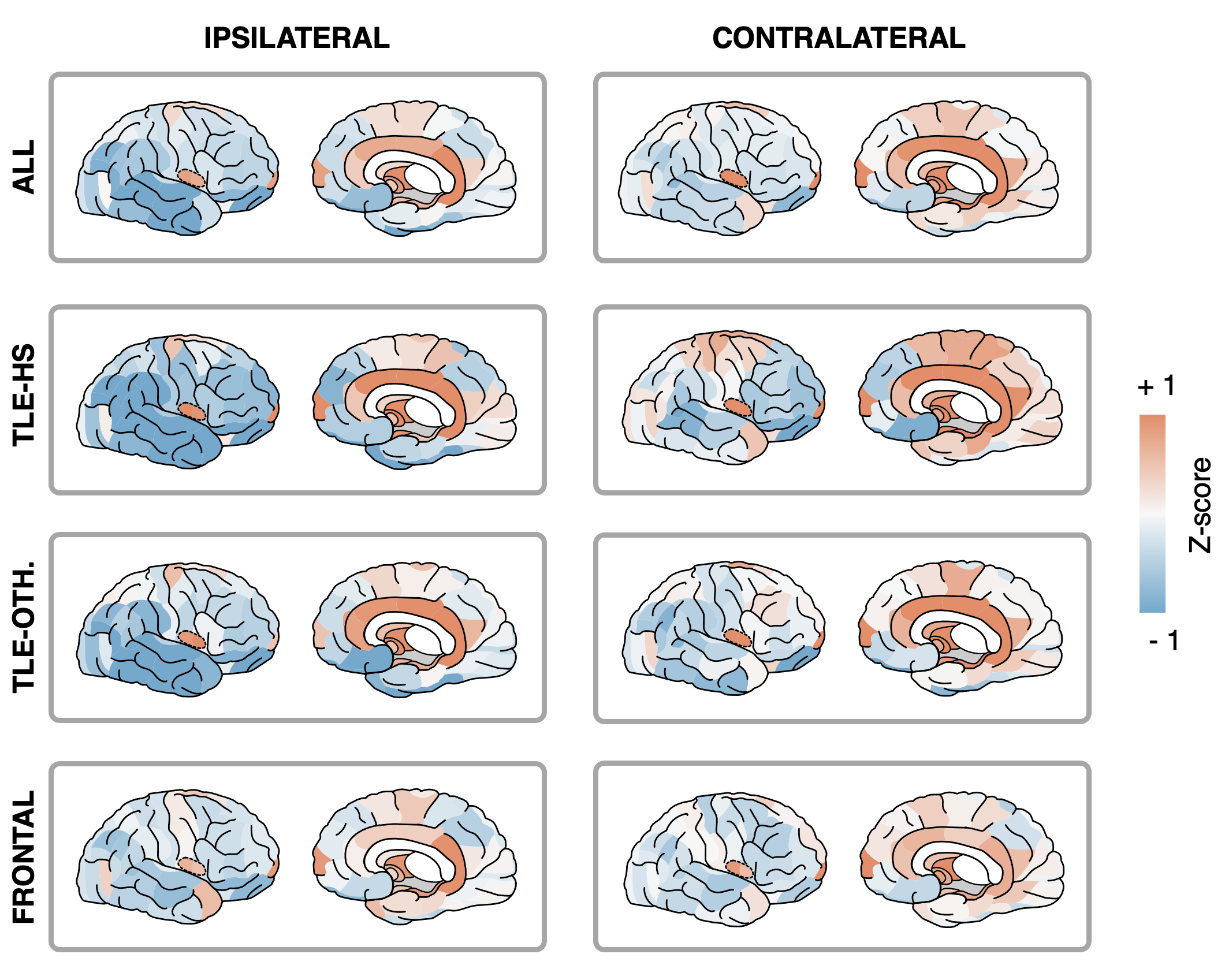

**Figure S3. Mean structural connectivity strength of each brain parcellation in patients with focal epilepsy.** Each region has a connectivity strength, which is the sum of all the SIFT2 score underling each its connections/edges. Values were corrected for age, sex, mean connectivity strength, total intracranial volume and subject motion and then Z-scored against the control group distribution. Subgroup data is provided for patients with temporal lobe epilepsy (TLE) with hippocampal sclerosis (TLE-HS), without HS (TLE-OTH.) and frontal epilepsy (FRONTAL). The columns provide the values of each brain region parcellations ipsilateral and contralateral to seizure onset zone, respectively.

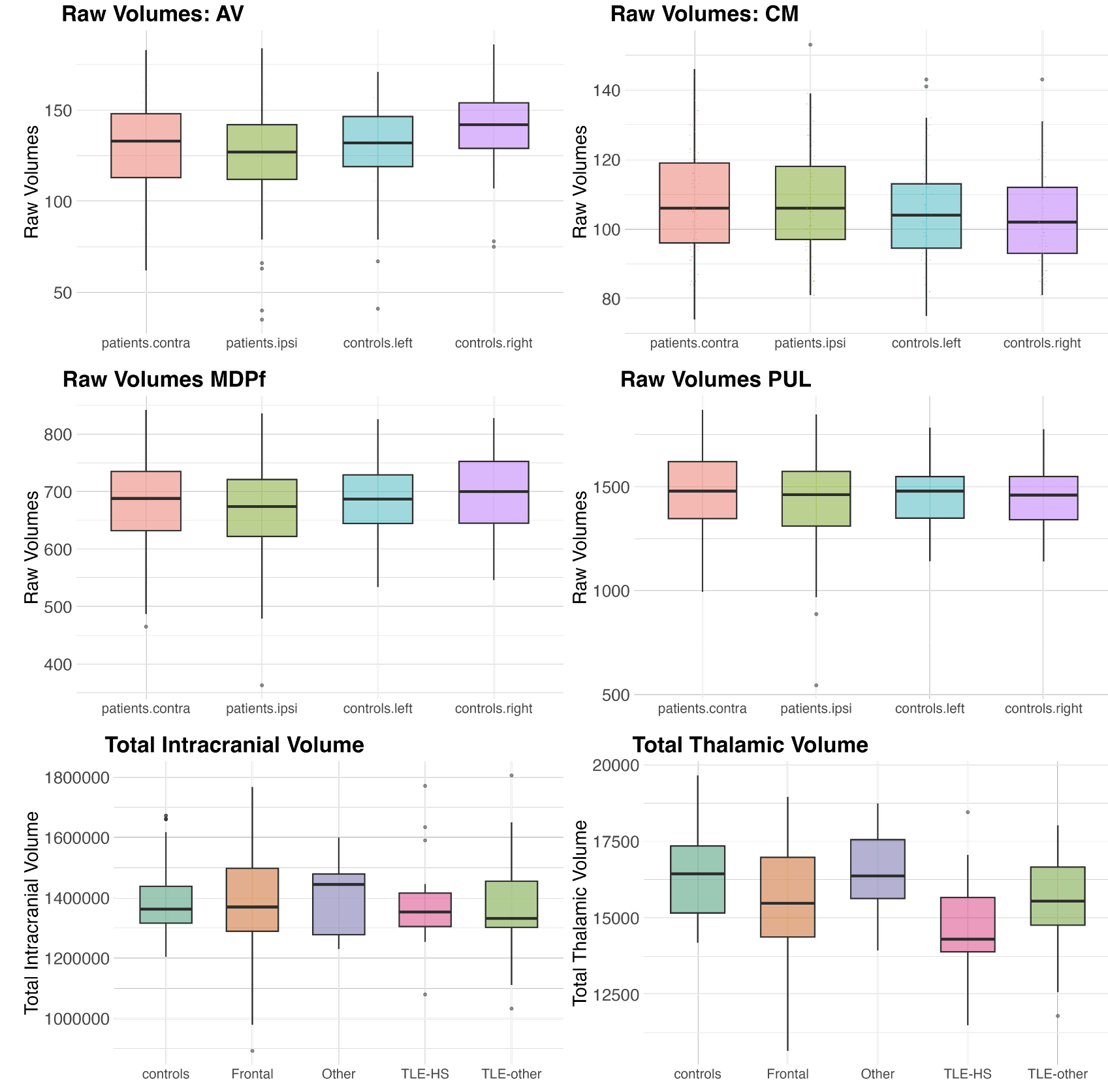

**Figure S4. Raw volumes of each thalamic nucleus, total intracranial volume and total thalamic volume.** Total thalamic volume is derived from the *Freesurfer* segmentation. AV = anteroventral nucleus of the thalamus; CM = centromedian nucleus of the thalamus; MDPf = mediodorsal-parafasicular nucleus of the thalamus; PUL = pulvinar nucleus of the thalamus.

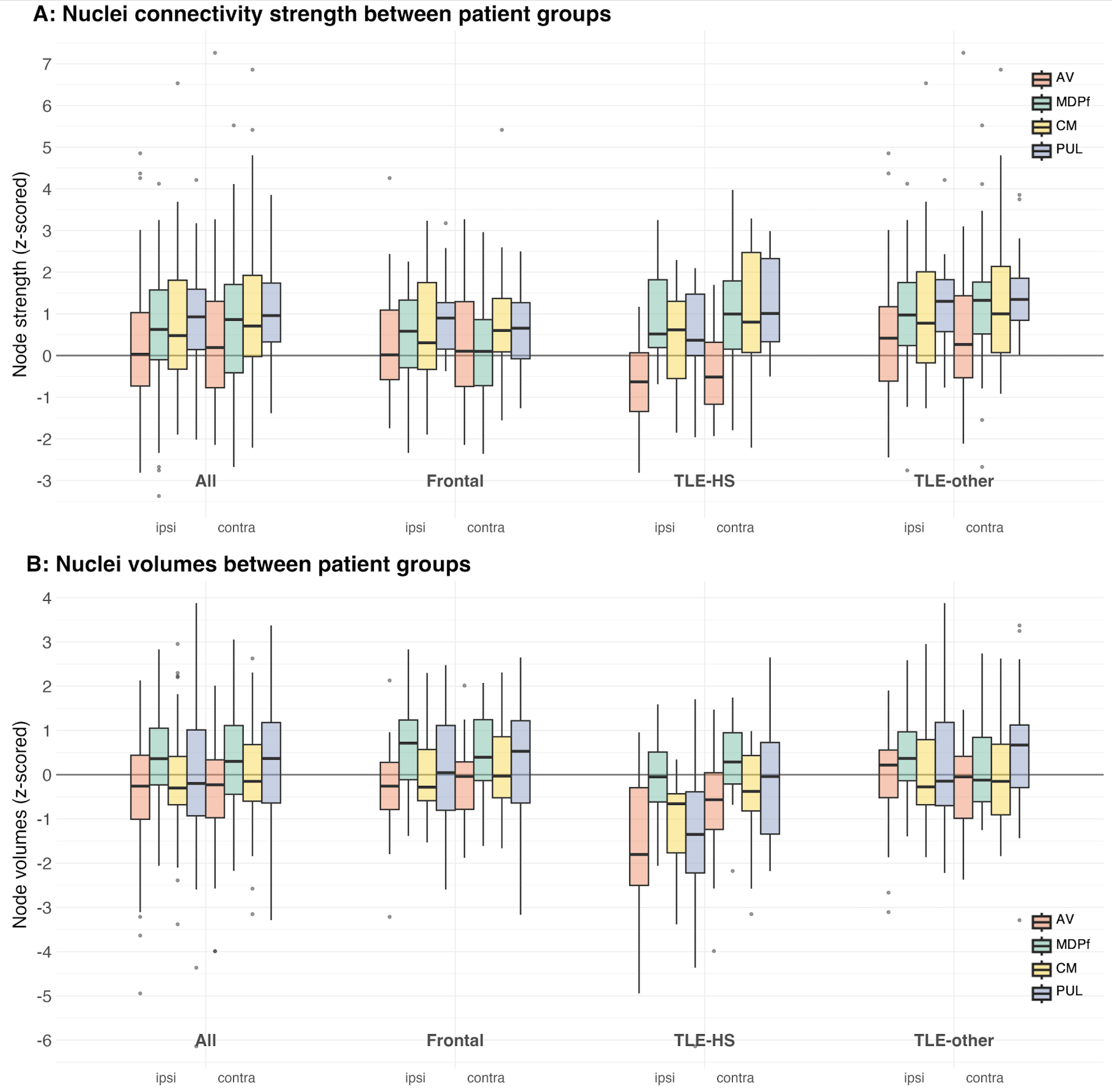

**Figure S5.** **Structural connectivity and** **volumetric signatures of the ipsilateral and contralateral thalamic nuclei in children with focal epilepsy. A:** Nuclei connectivity strength in all patient and subgroups of patients with temporal lobe epilepsy with hippocampal sclerosis (TLE-HS), temporal lobe epilepsy without hippocampal sclerosis (TLE-other), frontal seizure onset zones. All connectivity strengths are z-scored to control distributions after correction for age, sex, mean node connectivity strength, and motion. **B:** Nuclei volumes in the same cohorts. All volumes are z-scored to control distributions after correction for age, sex, and total intracranial volume. Plots show the median value and interquartile ranges. AV = anteroventral nucleus of the thalamus; CM = centromedian nucleus of the thalamus; MDPf = mediodorsal-parafasicular nucleus of the thalamus; PUL = pulvinar nucleus of the thalamus.

**Appendix 1.** **Nuclei connectivity strengths in patients vs. controls.** Ageneral linear model was used to model the structural connectivity scores of each the thalamic nuclei, using the variables: nucleus (AV, CM, MDPf, PUL), side (right or left), an group (controls and patients). *Pillau’s value* was used to report the multivariate tests and *Type III Sum of Squares* was used to report the between subject effects in the models.

| **Within-Subjects Factors** | | |
| --- | --- | --- |
| **Nucleus** | **Side** | **Dependent Variable** |
| AV | Left | AV Left |
|  | Right | AV Right |
| CM | Left | CM Left |
|  | Right | CM Right |
| MDPf | Left | MDPf Left |
|  | Right | MDPf Right |
| PUL | Left | PUL Left |
|  | Right | PUL Right |

| **Between-Subjects Factors** | | |
| --- | --- | --- |
|  | | **N** |
| **Group** | **Controls** | 63 |
|  | **Patients** | 81 |

| **Multivariate Tests^a^** | | | | | | | |
| --- | --- | --- | --- | --- | --- | --- | --- |
| **Effect** | **Pillai's Trace Value** | **F** | **Hypothesis df** | **Error df** | **Sig.** | **Partial Eta Squared (η²ₚ)** | **Observed Power** |
| **Nucleus** | **0.079** | **4.005** | **3.000** | **140.000** | **0.009** | **0.079** | **0.829** |
| **Nucleus * Group** | **0.079** | **4.005** | **3.000** | **140.000** | **0.009** | **0.079** | **0.829** |
| Side | 0.002 | 0.310 | 1.000 | 142.000 | 0.579 | 0.002 | 0.086 |
| Side * Group | 0.002 | 0.310 | 1.000 | 142.000 | 0.579 | 0.002 | 0.086 |
| Nucleus * Side | 0.023 | 1.108 | 3.000 | 140.000 | 0.348 | 0.023 | 0.294 |
| Nucleus * Side * Group | 0.023 | 1.108 | 3.000 | 140.000 | 0.348 | 0.023 | 0.294 |
| a. Design: Intercept + Group  Within Subjects Design: Nucleus + Side + Nucleus * Side | | | | | | | |

| **Tests of Between-Subjects Effects** | | | | | | | |
| --- | --- | --- | --- | --- | --- | --- | --- |
| **Source** | **Type III Sum of Squares** | **df** | **Mean Square** | **F** | **Sig.** | **Partial Eta Squared (η²ₚ)** | **Observed Power^a^** |
| Intercept | 133.799 | 1 | 133.799 | 24.325 | <0.001 | 0.146 | 0.998 |
| Group | **133.799** | **1** | **133.799** | **24.325** | **<0.001** | **0.146** | **0.998** |
| Error | 781.075 | 142 | 5.501 |  |  |  |  |

**Appendix 2.** **Nuclei connectivity volumes in patients vs. controls.** A general linear model was used to model the volumes of each the thalamic nuclei, using the variables: nucleus (AV, CM, MDPf, PUL), side (right or left), and group (controls and patients). *Pillau’s value* was used to report the multivariate tests and *Type III Sum of Squares* was used to report the between subject effects in the models.

| **Within-Subjects Factors** | | |
| --- | --- | --- |
| **Nucleus** | **Side** | **Dependent Variable** |
| AV | Left | AV_left |
|  | Right | AV_right |
| CM | Left | CM_left |
|  | Right | CM_right |
| MDPf | Left | MDPf_left |
|  | Right | MDPf_right |
| PULV | Left | PUL_left |
|  | Right | PUL_right |

| **Between-Subjects Factors** | | |
| --- | --- | --- |
|  | | **N** |
| **Group** | **Controls** | 63 |
|  | **Patients** | 81 |

| **Multivariate Tests** | | | | | | | |
| --- | --- | --- | --- | --- | --- | --- | --- |
| **Effect** | **Pillai's Trace Value** | **F** | **Hypothesis df** | **Error df** | **Sig.** | **Partial Eta Squared (η²ₚ)** | **Observed Power** |
| **Nucleus** | **0.115** | **6.051** | **3.000** | **140.000** | **<0.001** | **0.115** | **0.955** |
| **Nucleus * Group** | **0.115** | **6.051** | **3.000** | **140.000** | **<0.001** | **0.115** | **0.955** |
| Side | 0.009 | 1.265 | 1.000 | 142.000 | 0.263 | 0.009 | 0.201 |
| Side * Group | 0.009 | 1.265 | 1.000 | 142.000 | 0.263 | 0.009 | 0.201 |
| Nucleus * Side | 0.014 | 0.679 | 3.000 | 140.000 | 0.566 | 0.014 | 0.191 |
| Nucleus * Side * Group | 0.014 | 0.679 | 3.000 | 140.000 | 0.566 | 0.014 | 0.191 |
| Design: Intercept + Group  Within Subjects Design: Nucleus + Side + Nucleus * Side | | | | | | | |

| **Tests of Between-Subjects Effects** | | | | | | | |
| --- | --- | --- | --- | --- | --- | --- | --- |
| **Source** | **Type III Sum of Squares** | **df** | **Mean Square** | **F** | **Sig.** | **Partial Eta Squared (η²ₚ)** | **Observed Power** |
| Intercept | 0.007 | 1 | 0.007 | 0.002 | 0.968 | <0.000 | 0.050 |
| Group | 0.007 | 1 | 0.007 | 0.002 | 0.968 | <0.000 | 0.050 |
| Error | 606.190 | 142 | 4.269 |  |  |  |  |

**Appendix 3. General linear model was used to model the structural connectivity strengths of the thalamic nuclei between the patients with different seizure-onset category**. Three subgroups of patients were included: TLE-HS, TLE-other and frontal. *Pillau’s value* was used to report the multivariate tests and *Type III Sum of Squares* was used to report the between subject effects in the models. Power was computed using alpha = 0.05.

| **Within-Subjects Factors** | | |
| --- | --- | --- |
| **Nucleus** | **Laterality** | **Dependent Variable** |
| AV | Ipsilateral | AV ipsilateral |
|  | Contralateral | AV contralateral |
| CM | Ipsilateral | CM ipsilateral |
|  | Contralateral | CM contralateral |
| MDPf | Ipsilateral | MDPf ipsilateral |
|  | Contralateral | MDPf contralateral |
| PUL | Ipsilateral | PUL ipsilateral |
|  | Contralateral | PUL contralateral |

| **Between-Subjects Factors** | | |
| --- | --- | --- |
|  | | **N** |
| SOZ (seizure onset zone) category | Frontal | 29 |
|  | TLE-HS | 16 |
|  | TLE-other | 28 |
| Seizure Freedom Status | NSF | 30 |
|  | SF | 43 |

| **Effect** | **Pillai's Trace Value** | **F** | **Hypothesis df** | **Error df** | **Sig.** | **Partial Eta Squared (η²ₚ)** | **Observed Power** |
| --- | --- | --- | --- | --- | --- | --- | --- |
| **Nucleus** | **0.280** | **8.427** | **3.000** | **65.000** | **<0.001** | **0.280** | **0.991** |
| Nucleus * SOZ | 0.117 | 1.370 | 6.000 | 132.000 | 0.231 | 0.059 | 0.521 |
| Nucleus * Seizure Freedom | 0.001 | 0.013 | 3.000 | 65.000 | 0.998 | 0.001 | 0.052 |
| Nucleus * SOZ * Seizure Freedom | 0.016 | 0.181 | 6.000 | 132.000 | 0.982 | 0.008 | 0.095 |
| Laterality | 0.047 | 3.322 | 1.000 | 67.000 | 0.073 | 0.047 | 0.435 |
| **Laterality * SOZ** | **0.099** | **3.690** | **2.000** | **67.000** | **0.030** | **0.099** | **0.659** |
| **Laterality * Seizure Freedom** | **0.109** | **8.176** | **1.000** | **67.000** | **0.006** | **0.109** | **0.805** |
| Laterality * SOZ * Seizure Freedom | 0.002 | 0.060 | 2.000 | 67.000 | 0.942 | 0.002 | 0.059 |
| Nucleus * Laterality | 0.061 | 1.403 | 3.000 | 65.000 | 0.250 | 0.061 | 0.356 |
| Nucleus * Laterality * SOZ | 0.101 | 1.164 | 6.000 | 132.000 | 0.329 | 0.050 | 0.446 |
| Nucleus * Laterality * Seizure Freedom | 0.082 | 1.932 | 3.000 | 65.000 | 0.133 | 0.082 | 0.477 |
| Nucleus * Laterality * SOZ * Seizure Freedom | 0.057 | 0.648 | 6.000 | 132.000 | 0.692 | 0.029 | 0.251 |

| **Tests of Between-Subjects Effects** | | | | | | | |
| --- | --- | --- | --- | --- | --- | --- | --- |
| **Source** | **Type III Sum of Squares** | **df** | **Mean Square** | **F** | **Sig.** | **Partial Eta Squared(η²ₚ)** | **Observed Power** |
| Intercept | 206.753 | 1 | 206.753 | 39.164 | <.001 | 0.369 | 1.000 |
| SOZ | 24.574 | 2 | 12.287 | 2.327 | 0.105 | 0.065 | 0.456 |
| Seizure Freedom | 2.452 | 1 | 2.452 | 0.464 | 0.498 | 0.007 | 0.103 |
| SOZ * Seizure Freedom | 10.522 | 2 | 5.261 | 0.997 | 0.375 | 0.029 | 0.217 |
| Error | 353.702 | 67 | 5.279 |  |  |  |  |

| **Multiple Comparisons** | | | | | | | |
| --- | --- | --- | --- | --- | --- | --- | --- |
|  | | **SOZ** | **Mean Difference (I-J)** | **Std. Error** | **Sig.** | **95% Confidence Interval** | |
|  |  |  |  |  |  | **Lower Bound** | **Upper Bound** |
| **SOZ** | **Frontal** | TLE-HS | -0.002 | 0.253 | 0.993 | -0.507 | 0.503 |
|  |  | **TLE-other** | **-0.489** | **0.215** | **0.026** | **-0.919** | **-0.059** |
|  | **TLE-HS** | Frontal | 0.002 | 0.253 | 0.993 | -0.503 | 0.507 |
|  |  | TLE-other | -0.487 | 0.255 | 0.060 | -0.995 | 0.021 |
|  | **TLE-other** | **Frontal** | **0.489** | **0.215** | **0.026** | **0.0594** | **0.919** |
|  |  | TLE-HS | 0.487 | 0.255 | 0.060 | -0.0215 | 0.995 |

**Appendix 4.** **General linear model was used to model the volumes of the thalamic nuclei between the patients with different seizure-onset category**. Three subgroups of patients were included: TLE-HS, TLE-other and frontal. *Pillau’s value* was used to report the multivariate tests and *Type III Sum of Squares* was used to report the between subject effects in the models. Power was computed using alpha = 0.05.

| **Within-Subjects Factors** | | |
| --- | --- | --- |
| **Nucleus** | **Laterality** | **Dependent Variable** |
| AV | Ipsilateral | AV ipsilateral |
|  | Contralateral | AV contralateral |
| CM | Ipsilateral | CM ipsilateral |
|  | Contralateral | CM contralateral |
| MDPf | Ipsilateral | MDPf ipsilateral |
|  | Contralateral | MDPf contralateral |
| PUL | Ipsilateral | PUL ipsilateral |
|  | Contralateral | PUL contralateral |

| **Between-Subjects Factors** | | |
| --- | --- | --- |
|  | | **N** |
| SOZ (seizure onset zone) category | Frontal | 29 |
|  | TLE-HS | 16 |
|  | TLE-other | 28 |
| Seizure Freedom Status | NSF | 30 |
|  | SF | 43 |

| **Multivariate Tests** | | | | | | | |
| --- | --- | --- | --- | --- | --- | --- | --- |
| **Effect** | **Pillai's Trace Value** | **F** | **Hypothesis df** | **Error df** | **Sig.** | **Partial Eta Squared (η²ₚ)** | **Observed Power** |
| **Nucleus** | **0.374** | **12.918** | **3.000** | **65.000** | **<0.001** | **0.374** | **1.000** |
| Nucleus * SOZ | 0.128 | 1.508 | 6.000 | 132.000 | 0.180 | 0.064 | 0.568 |
| Nucleus * Seizure Freedom | 0.013 | 0.281 | 3.000 | 65.000 | 0.839 | 0.013 | 0.101 |
| Nucleus * SOZ * Seizure Freedom | 0.068 | 0.779 | 6.000 | 132.000 | 0.588 | 0.034 | 0.300 |
| **Laterality** | **0.110** | **8.296** | **1.000** | **67.000** | **0.005** | **0.110** | **0.810** |
| **Laterality * SOZ** | **0.262** | **11.873** | **2.000** | **67.000** | **<0.001** | **0.262** | **0.993** |
| **Laterality * Seizure Freedom** | **0.073** | **5.237** | **1.000** | **67.000** | **0.025** | **0.073** | **0.616** |
| Laterality * SOZ * Seizure Freedom | 0.055 | 1.954 | 2.000 | 67.000 | 0.150 | 0.055 | 0.391 |
| **Nucleus * Laterality** | **0.268** | **7.952** | **3.000** | **65.000** | **<0.001** | **0.268** | **0.987** |
| **Nucleus * Laterality * SOZ** | **0.185** | **2.245** | **6.000** | **132.000** | **0.043** | **0.093** | **0.772** |
| Nucleus * Laterality * Seizure Freedom | 0.073 | 1.705 | 3.000 | 65.000 | 0.175 | 0.073 | 0.426 |
| Nucleus * Laterality * SOZ * Seizure Freedom | 0.058 | 0.656 | 6.000 | 132.000 | 0.685 | 0.029 | 0.254 |
| Design: Intercept + SOZ + Seizure Freedom + SOZ * Seizure Freedom  Within Subjects Design: Nucleus + Laterality + Nucleus * Laterality | | | | | | | |

| **Tests of Between-Subjects Effects** | | | | | | | |
| --- | --- | --- | --- | --- | --- | --- | --- |
| **Source** | **Type III Sum of Squares** | **df** | **Mean Square** | **F** | **Sig.** | **Partial Eta Squared (η²ₚ)** | **Observed Power** |
| Intercept | 8.536 | 1 | 8.536 | 1.777 | 0.187 | 0.026 | 0.260 |
| **SOZ** | **47.727** | **2** | **23.863** | **4.967** | **0.010** | **0.129** | **0.794** |
| Seizure Freedom | 0.334 | 1 | 0.334 | 0.069 | 0.793 | 0.001 | 0.058 |
| SOZ * Seizure Freedom | 0.909 | 2 | 0.455 | 0.095 | 0.910 | 0.003 | 0.064 |
| Error | 321.923 | 67 | 4.805 |  |  |  |  |

| **Multiple Comparisons** | | | | | | |
| --- | --- | --- | --- | --- | --- | --- |
| **SOZ** | **SOZ** | **Mean Difference** | **Std. Error** | **Sig.** | **95% Confidence Interval** | |
|  |  |  |  |  | **Lower Bound** | **Upper Bound** |
| Frontal | **TLE-HS** | **0.77** | **0.24** | **0.002** | 0.29 | 1.25 |
|  | TLE-other | 0.08 | 0.21 | 0.696 | -0.33 | 0.49 |
| TLE-HS | **Frontal** | **-0.77** | **0.24** | **0.002** | -1.25 | -0.29 |
|  | **TLE-other** | **-0.69** | **0.24** | **0.006** | -1.18 | -0.21 |
| TLE-other | Frontal | -0.08 | 0.21 | 0.696 | -0.49 | 0.33 |
|  | **TLE-HS** | **0.69** | **0.24** | **0.006** | 0.21 | 1.18 |
