## Supplementary material for "Thalamocortical structural connectivity in children with focal epilepsy: a diffusion MRI, case-control study": Table 1

**Table 1. Demographics and clinical details of the control and patient cohorts.** IQR = inter-quartile range; F = female; M = male; TLE = temporal lobe epilepsy: HS = hippocampal sclerosis; FBTCS = focal-to-bilateral tonic-clonic seizures. *Not accounting for missing data.

|  | **Controls** | **Patients (all)** | **Patients: frontal** | **Patients: TLE-HS** | **Patients: TLE-other** | **Patients: other** |
| --- | --- | --- | --- | --- | --- | --- |
| **Demographics** |  |  |  |  |  |  |
| Number of children | 63 | 81 | 29 | 16 | 29 | 7 |
| Age at MRI scan  (median, IQR) in years | 12.8,  (9.6 - 12.5) | 12.2,  (9.6-16.0) | 10.9 (9.5-14.1) | 15.0 (12.5-17.6) | 12.1 (9.7-16.0) | 10.6, (8.6-11.9) |
| Sex (F:M ratio) | 49:14 | 40:41 | 14:15 | 10:6 | 13:16 | 3:4 |
| Follow-up duration (median, IQR) in years | - | 1.7 (1.2-2.8) | 1.6 (1.2-3.6) | 2.0 (1.3 – 2.3) | 1.5 (1.2-2.4) | 2.4 (1.8-3.4) |
| Duration of epilepsy (median, IQR) in years * | - | 5.4 (3.5-8.7) | 4.8 (2.5-8.4) | 9.4 (4.0-15.1) | 5.4 (5.8-8.4) | 4.1 (3.9-5.9) |
| **Resection cavities** |  |  |  |  |  |  |
| Right : Left | - | 32:49 | 11:18 | 8:8 | 17:12 | 1:6 |
| **Histopathology** |  |  |  |  |  |  |
| Focal cortical dysplasia | - | 23 | 15 | 0 | 6 | 2 |
| Tumours | - | 24 | 6 | 0 | 17 | 1 |
| Hippocampal sclerosis | - | 16 | 0 | 16 | 0 | 0 |
| Other | - | 18 | 8 | 0 | 6 | 4 |
| **Postop. seizure outcome** |  |  |  |  |  |  |
| Seizure Free | - | 47 | 19 | 6 | 18 | 4 |
| Not Seizure Free | - | 32 | 10 | 10 | 10 | 2 |
| Unknown | - | 2 | 0 | 0 | 1 | 1 |
| **Seizure types** |  |  |  |  |  |  |
| FBTCS | - | 31 | 9 | 9 | 11 | 2 |
| No FBTCS | - | 44 | 17 | 5 | 17 | 5 |
| Unknown | - | 6 | 3 | 2 | 1 | 0 |
